# KidneyChain: Leveraging Blockchain & Artificial Intelligence for a Streamlined Organ Donation Solution

**DOI:** 10.1101/2024.06.08.24308145

**Authors:** Kapil Panda, Anirudh Mazumder

## Abstract

Currently the kidney organ transplantation process is a manual and cumbersome process, resulting in organ wastage and reoccurring issues in patients due to a lack of organ health screening prior to transplantation. To solve these issues in the industry, we utilize blockchain technology and artificial intelligence to create a streamlined kidney transplantation process. Utilizing blockchain technology, we automated the organ matching process, and matched donors and patients based on the same metrics that UNOS clinicians use, resulting in efficient and faster matching. Furthermore, using artificial intelligence, we created a model with perfect accuracy that screens organs pre-transplantation to predict possible risk for common kidney diseases such as chronic kidney disease(CKD), acute kidney injury(AKI), and polycystic kidney disease(PKD), to prevent recurring issues in patients, post-transplantation. Furthermore, a tapped delay line convolutional neural network provides cybersecurity by identifying valid blockchain transactions from fake patterns with 100% accuracy, ensuring full data privacy. By synergizing blockchain, AI, and cybersecurity, this research creates an efficient, secure platform that could expand patient access to life-saving transplants, prevent transplant failures, and save thousands of lives currently lost due to inefficiencies and wait times.

## 1. Introduction

Kidney transplants are widely viewed as the best treatment for end-stage renal disease and kidney failure, offering patients the chance to resume a healthy, normal lifestyle, however, every day 12 people die waiting for a transplant with more than 100,000 people in the US alone on the transplant waitlist [1]. Currently in the organ donation system, the United Network for Organ Sharing(UNOS) manually matches donors to patients, however, due to the various factors that that play a role in organ matching, such as blood type, HLA, hemoglobin levels, etc., this process is prolonged and highly inefficient, contributing to over 28,000 organs being wasted and several thousand deaths from patients on the transplant waitlist [2]. Furthermore, the kidney transplantation process specifically lacks a coherent way to screen organ health before transplantation to determine future risk for kidney disease, resulting in 30% of patients with kidney transplants having recurring issues. In fact this issue has become so prevalent that it has even required Congress to intervene with the recent passing of the Organ Procurement and Transplantation Network Modernization Initiative to foster innovation in this space [3].

To solve these issues in the industry, we utilize blockchain technology and artificial intelligence to create a streamlined kidney transplantation process. Utilizing blockchain technology, we can automate the organ matching process and match donors and patients based on the same metrics that UNOS clinicians use, resulting in efficient and faster matching.

## 2. Methodology

In the following study, there were three components used to create the platform: blockchain, artificial intelligence, and cybersecurity as seen in Figure 1:

**Figure 1.**
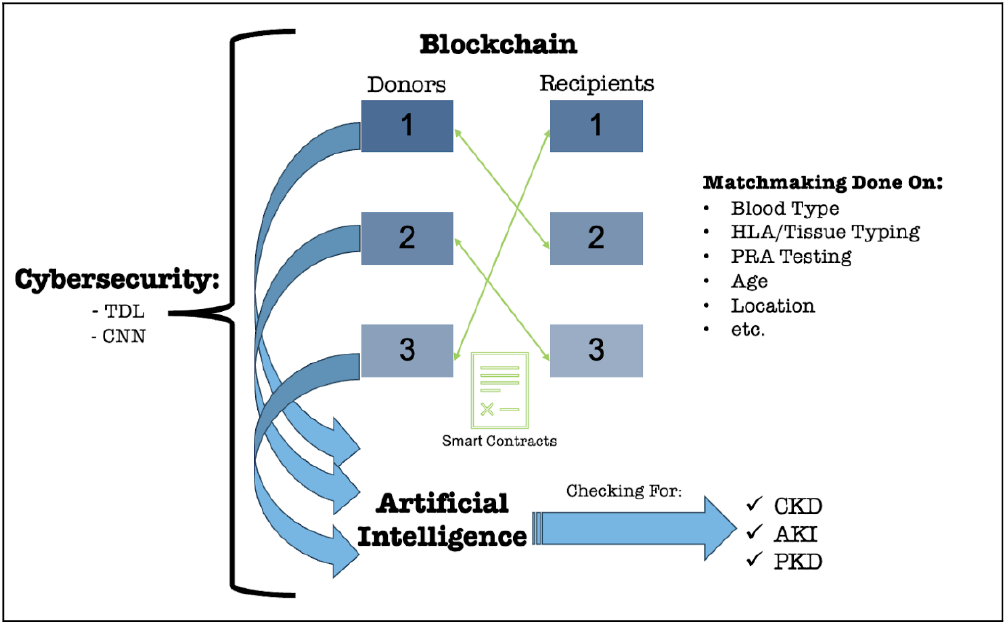
KidneyChain Platform Depiction

### 2.1. Blockchain

In this study, we employ blockchain technology to improve the organ transplantation process. The central goal of implementing blockchain in this project is to create a decentralized and secure platform [4] that automates the organ matching process, addressing the inefficiencies and shortcomings of the existing manual system.

KidneyChain targets donors and recipients at the same time. In KidneyChain’s network, we utilize the same data for either type of consumer(name, age, location, blood type, tissue type, PRA testing, etc.). However, we modified the conceptual blockchain to fit our needs and connect patients in-need of new kidneys efficiently [5]. First, we utilize two different chains: one for donors and one for recipients. This allows us to keep a organized but separated list of different patients that can be connected through KidneyChain. By keeping two separated lists, we can easily merge two types of patients into entire chain where managing the additions and pairings of each ‘sub-chain’ (one for donors and can be done.

In the blockchain-based matching system, the process begins with the input of donor and patient data into the secure and decentralized network. This data includes vital information that UNOS uses to match donors and patients such as blood type, human leukocyte antigen (HLA) markers, organ size, hemoglobin levels, tissue typing, PRA testing, age, location, wait time, CPRA score, and pediatric status [5]. The key to the matching process is the use of smart contracts, which are autonomous and optimizing pieces of code that contain predefined criteria and algorithms for evaluating donor-patient pairs. These smart contracts mimic the decision-making process of experienced healthcare professionals at UNOS, as they analyze the data for each potential pair and assign a compatibility score based on how well they meet the specified criteria. The criteria used within these smart contracts are designed to reflect the standards and parameters employed by traditional manual matchmakers. Once the smart contracts have assessed the potential donor-patient pairs, they rank these pairs based on their compatibility scores [6]. Higher-ranked pairs are indicative of a stronger match according to the defined criteria. This ranking system enables a systematic and efficient approach to organ matching, greatly reducing the time patients spend on waiting lists. The blockchain’s transparency and traceability are crucial aspects of this process. Every step of the organ matching process, from the input of data to the final matching decision, is recorded on the blockchain’s distributed ledger. This ensures that the entire process can be verified and audited, eliminating potential human errors and ensuring the integrity of the matching decisions. The blockchain’s immutability guarantees that the recorded data remains secure and tamper-proof.

#### 2.1.1. Hashing and Genesis Block Creation

For the functionality of KidneyChain’s network, we utilize a modified version of a regular blockchain network. In a regular blockchain network, we have a chain that is linked through previous hashes. The first block is normally established before deployment of the blockchain, commonly referred to as the genesis block. The genesis block contains 5 major values: the timestamp of when the block was created, the data the block contains, the nonce value, the current hash, and the previous hash [7].

The timestamp of each block is calculated using the ‘seconds-after-epoch’ value, which is simply the seconds after the date 01/01/1970. We can use the timestamp to relate the chronological order of blocks when measuring relevant values or for a simple stamp to identify each block. The data is any type of data that can be stored in a block. One important benefit of a blockchain network is the variability of data that can potentially be stored within the block: transactions, monetary values, and more. In our case, we will use a JSON-formatted data object for each block. JSON is a shorthand-notation for ‘JavaScript Object Notation’ which is resemblent a dictionary, having key-value pairs within each JSON object. The nonce value is a value which is shorthand-notation for ‘number only created once,’ which is important in mining for blockchain. Mining is the process of using complex mathematical processes and high computational power to ‘discover’ new blocks for the chain, but, as we see, this feature becomes less important to our scenario. The current hash is a ‘human fingerprint’ for each block, identifying each block with a unique identifier nearly impossible to replicate. This fingerprint is a hash value which can be represented in many different ways, based on the specific hash function used (which range from least to most secure). One of the most common but most secure hash functions is SHA-256, named after its generated of 256 bit or 64 character length hash values with hash values being represented in hexadecimal values. In terms of security, the length of a SHA-256 generated value being repeated for two different inputs is 2^256^, making it nearly impossible for two different blocks to have the same identity. The previous hash is how blocks are connected and a chain is formed, hence the name ‘blockchain.’ Each block excluding the genesis block contains a previous hash value which is simply the generated hash value for the previous block or the ‘identity’ of the previous block [7].

#### 2.1.2. Matching Algorithm in Solidity

The final part of a blockchain is the Smart Contract or individual pieces of code that are attached to the entire blockchain and distributed to each block within. A smart contract simply states a set of instructions within code that informs what each block should do in case of a certain scenario such as a transaction or a hash value generation. In other words, a smart contract acts as an automated middleman within a blockchain. Smart contracts are normally created using blockchain programming language with the most popular being Solidity from the cryptocurrency Ethereum: An easy-to-use Turing-complete (ability for logic to be implemented) programming language for creating automated switches in a blockchain [6].

While Smart Contracts are Turing-complete, they still cannot create or deploy Machine Learning models, hindering the purpose of including the proposed matching algorithm to the blockchain. However, KidneyChain deploys the proposed matching algorithm using internal API services on Python using Flask. To utilize this API, the Solidity contract uses oracles – third-party services in Solidity. More specifically, we use ChainLink which is an established API service for Solidity with byte addresses and other valuable information. We utilize ChainLink and connect to our deployed ML model, feeding our information such as age, PRA testing, and more, to the API. We get back the results from the API and we can perform ML-based donor-to-patient matching on Solidity.

After we implement the contract in Solidity, we still have to achieve a connection between the blockchain in Python with the contract – this is done through Web3 technologies. First, we compile our Solidity contract by converting it to a unique binary or bytecode. We do this through an external ‘solc’ library which provides easy compilation of the contract. Next, we use a Web3 Deployment API where we can ‘host’ our Solidity contract for connection with a blockchain. Finally, we connect the blockchain in Python using an external ‘web3’ package, where we can load and deploy the smart contract to our blockchain system and modulize each block.

#### 2.1.3. Inherent Security Measures and Defense Protocol

Besides secure hash values and complex chains, there are more security measures that a regular blockchain takes and ones we incorporate in KidneyChain.

First, blockchain utilizes an immutable ledger system. Resemblance to deeds in a housing market, there must exist a tampering-proof ledger system that prevents people from tampering with blocks in a blockchain. An immutable ledger system simply instates that blocks or smart contracts (although created on a separate basis) CANNOT be changed once deployed. This principle is more important than a regular housing system, however, since changing one block in a blockchain changes every block afterward (each block contains a previous hash).

Second, blockchain utilizes a distributed P2P network system, or a decentralized system to control additions and changes with blocks in the chain. Suppose faulty data in a block was attempted to be added to the chain. In that case, we do not have to worry about having one central authority to validate it: different copies of the blockchain from different middlemen and access points (different nodes in the network) can collectively detect this and instantly reject the addition of the new block. With a P2P network, the difficulty of intentionally tampering with or accidentally adding data to the blockchain becomes exponentially harder with an immutable ledger system in place as well [7].

### 2.2. Cybersecurity

Although blockchain has its own measures to ensure data security through cryptography and decentralized consensus, additional cybersecurity methods are still needed to fully secure the system, given the sensitive medical data transmitted across the network. To augment the blockchain’s innate security, this platform incorporates an advanced artificial intelligence cybersecurity module. This provides real-time threat detection by using deep learning to analyze patterns in the blockchain data and distinguish valid from invalid transactions11. Specifically, a novel tapped delay line convolutional neural network (TDL-CNN) was implemented for this role [8]

A TDL-CNN model revolves around the importance of a single input vector consisting of time-series values with a specified number of timesteps. This time-series data revolves around one specific unit variable, which is what we are testing. Mathematically, the tapped-delay line of the TDL-CNN can be represented as

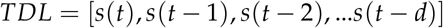

where *t* is the current timestep and *d* is the delay or the furthest point to look back to.

Additionally, one statement must be true for the TDL-CNN to work in simulation. For a delay *d* and total dataset size of *N, d* < *N* for any point in the TDL-CNN training or testing process or there is insufficient data for the TDL-CNN to work.

Finally, for the actual prediction of the model, the CNN predicts *s*(*t* + 1) using the input vector provided. When predicting using the input vector, the prediction gets pushed onto the unit vector as

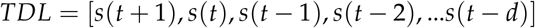

and the TDL-CNN moves on the next timestep until *t* >= *N* [9].

## 3. Results

### 3.1. Blockchain

The proposed blockchain representation is functional with its representation in blockchain Not only does the proposed JSON-formatted blockchain provide a safe, accessible data storage system but it is also able to be represented visually using HTML and JavaScript itself, ensuring that the proposed user interaction is sufficient. However, the smart contract has been implemented with a mix of Solidity and Python instead of completely Solidity. We believe that more external libraries such as an external Solidity function like AggregatorV3Interface for API interaction could provide more results. However, the matching algorithm has demonstrated its applicability and functionality within the blockchain, as all the proposed features are used and applied in the matching algorithm. Another limitation is the modified distributed P2P network as the concept of registering node addresses and checks needs to be changed specifically – the P2P network can be researched more. Fortunately, the general area of security within the blockchain regarding immutable ledgers works completely fine as security protocols don’t allow any modifications after hashing to be made.

## 4. Discussion

This research demonstrates the immense potential for emerging technologies like blockchain, artificial intelligence, and cybersecurity to drive transformative change in organ transplantation. The proposed KidneyChain platform offers a glimpse into how thoughtfully integrating these innovations could reshape and optimize complex healthcare processes. This work carries major implications for the future of organ matching, transplantation outcomes, and even human lives [10]. The reasoning behind implementing blockchain technology is clear - to bring enhanced transparency, decentralization, automation, and security to the organ procurement process. By encoding expert matching criteria into smart contracts and recording all transactions on a tamper-proof ledger, blockchain removes human error risks and makes the system auditable. This research shows blockchain’s viability for managing sensitive medical data and executing rules-based healthcare processes.

Together, these innovations stand to significantly expand patient access to transplants, prevent wasted organs, and save thousands of lives annually. Broader deployment would generate richer datasets to refine the smart contracts, improve predictive algorithms, and strengthen security protections. This work positions blockchain, AI, and cybersecurity at the forefront of modernizing critical healthcare infrastructure. These technologies undoubtedly face regulatory, ethical, and adoption challenges. Thoughtful design and testing will be imperative before real-world implementation. But the potentials are enormous - well beyond transforming organ transplantation. The concepts demonstrated here could be extended to optimize allocation, monitoring, and maintenance of precious healthcare resources. And the synergistic integration of blockchain, AI, and cybersecurity provides a model for digitally transforming many complex industries and processes.

By proving these emerging technologies’ viability and value for life-saving medical applications, this research sparks hope for the future possibilities of technological innovation to profoundly improve human lives and society. KidneyChain offers just one glimpse into the transformative potential of thoughtfully combining blockchain, AI, and cybersecurity to drive positive change.

## 5. Conclusions

This research demonstrates the potential for blockchain technology and artificial intelligence to transform and improve the kidney transplantation process. The blockchain-based organ matching system leverages smart contracts to replicate manual decision-making in an automated, decentralized manner, enabling faster and more efficient donor-patient pairing. By recording all transactions transparently on an immutable ledger, the system also ensures full traceability and auditing capabilities. Together, these innovations create a comprehensive platform that connects organ procurement organizations, transplant centers, and patients through enhanced transparency, security, and automation. By synergizing emerging technologies, this system has the potential to expand access to transplants, prevent wasted organs, and ultimately save thousands of lives lost annually due to inefficiencies and delays. This research provides a prototype for blockchain and AI’s ability to modernize and optimize complex healthcare processes. Further development and testing in real-world settings could help drive adoption and validate the effectiveness of the proposed platform. There remain regulatory and implementation challenges, but solutions like KidneyChain demonstrate the immense opportunity for technological innovation to reshape the organ transplantation landscape.

## 6. Future Work

Future research should focus on expanding the dataset and model capabilities, conducting pilot studies for clinical validation, enhancing security protections, improving usability through user-centered design, and investigating alternative distributed ledger architectures. By iteratively refining the proposed technologies through additional research along these lines, this work could eventually help realize the immense potentials of blockchain, artificial intelligence, and cybersecurity innovations to save lives through transformation of the organ transplant system. With rigorous ongoing research and development, the solutions pioneered here could evolve into robust and deployable platforms that demonstrably improve outcomes for patients awaiting life-saving transplants [11].

## 7. Patents

Patent Filed:

Name- KidneyChain

Number- US63596309

Date- November 6, 2023

## Data Availability

All data produced in the present work are contained in the manuscript.

## Acknowledgments

We would like to thank the University of North Texas for providing us with the resources and support to conduct this research. The invaluable guidance and encouragement from our professors and mentors have been instrumental in shaping the direction and scope of this study. We would also like to acknowledge the National Kidney Foundation for their support and inspiration in conducting this project. Finally, we would also like to thank our families for supporting us throughout our research.

## Disclaimer/Publisher’s Note

The statements, opinions and data contained in all publications are solely those of the individual author(s) and contributor(s) and not of MDPI and/or the editor(s). MDPI and/or the editor(s) disclaim responsibility for any injury to people or property resulting from any ideas, methods, instructions or products referred to in the content.

## References

1. Organ Donation Statistics. https://www.organdonor.gov/learn/organ-donation-statistics/, 2024. [Online; accessed 05-April-2008].

2. How we match organs. https://unos.org/transplant/how-we-match-organs/, 2024. [Online; accessed 04-April-2024].

3. Organ Procurement and Transplantation Network (OPTN) Modernization Initiative. https://www.hrsa.gov/optn-modernization/, 2024. [Online; accessed 04-March-2024].

4. Levis, D.; Fontana, F.; Ughetto, E. A look into the future of blockchain technology. PLoS ONE 2021, 16. 10.1371/journal.pone.0258995.

5. Ekblaw, A.; Azaria, A.; Halamka, J.D.; Lippman, A.; Vieira, T. A Case Study for Blockchain in Healthcare: “MedRec” prototype for electronic health records and medical research data White Paper MedRec: Using Blockchain for Medical Data Access and Permission Management IEEE Original Authors, 2016.

6. Azaria, A.; Ekblaw, A.; Vieira, T.; Lippman, A. MedRec: Using Blockchain for Medical Data Access and Permission Management. In Proceedings of the 2016 2nd International Conference on Open and Big Data (OBD), 2016, pp. 25–30. 10.1109/OBD.2016.11.

7. Nakamoto, S. Bitcoin: A peer-to-peer electronic cash system 2008.

8. Coutinho, F.D.; Silva, H.S.; Georgieva, P.; Oliveira, A.S. CNN Architecture Design Impact in DL-Based Channel Estimation Algorithms. In Proceedings of the 2023 Workshop on Communication Networks and Power Systems (WCNPS), 2023, pp. 1–7. 10.1109/WCNPS60622.2023.10345278.

9. Bakalos, N.; Voulodimos, A.; Doulamis, N.; Doulamis, A.; Ostfeld, A.; Salomons, E.; Caubet, J.; Jimenez, V.; Li, P. Protecting Water Infrastructure From Cyber and Physical Threats: Using Multimodal Data Fusion and Adaptive Deep Learning to Monitor Critical Systems. IEEE Signal Processing Magazine 2019, 36, 36–48. 10.1109/MSP.2018.2885359.

10. Systematic review: Kidney transplantation compared with dialysis in clinically relevant outcomes. American Journal of Transplantation 2011, 11, 2093–2109. 10.1111/j.1600-6143.2011.03686.x.

11. Raghupathi, W.; Raghupathi, V. Big data analytics in healthcare: promise and potential, 2014.

